# The bystander effect: lesion spatial extent as a foundational artifact in Lesion Network Mapping

**DOI:** 10.64898/2026.10.17.26365701

**Authors:** Shahar Edelman, Uri Elias, Shahar Arzy

## Abstract

Lesion network mapping (LNM) has provided a compelling framework in clinical neuroscience, linking focal brain damage at heterogeneous anatomical locations to shared functional networks across behavioural syndromes. Recently, the methodological specificity of LNM has come under discussion following observations that derived network maps tend to converge onto the normative connectome’s generic degree distribution. While current efforts largely treat this convergence as a statistical inference challenge addressable via post-hoc spatial null models or individual-level analyses, we identify a foundational upstream measurement artifact operating within individual patients: the lesion “bystander effect”. Because pathological lesions follow biological constraints rather than functional brain parcellations, a typical lesion encapsulates both a symptom-driving critical node and surrounding asymptomatic bystander tissue. By averaging connectivity profiles across the entire lesion mask, LNM mechanically dilutes focal clinical signals. Across empirical clinical cohorts, individual-level machine learning classifiers, synthetic within-cohort lesion unions, and whole-brain spherical simulations, we demonstrate that increasing lesion extent deterministically drives network maps toward the connectome degree vector following a mathematical power law. Fulfilling the clinical promise of LNM will therefore require computational strategies that isolate the causal functional core from surrounding bystander tissue prior to network projection.

## Introduction

Lesion network mapping (LNM) presented a transformative advance in clinical neuroscience. By utilizing large-scale normative functional connectomes to link structurally disparate lesions to shared neural circuits, LNM elegantly solved a long-standing localization paradox: how distinct lesioned loci cause similar symptoms.^1^ However, as the clinical applications of this powerful framework continue to expand,^2,3^ its methodological specificity was recently questioned. Van den Heuvel et al.^4^ demonstrated that LNM-derived circuits systematically collapse onto the connectome’s generic degree distribution. Initially framed as a tractable problem of group-level statistical inference, this observation has sparked constructive methodological debate. For instance, Zalesky and Cash^5^ proposed permutation-based spatial and topological null models, alongside a shift toward individual-level analyses, to better isolate disease signals from generic connectome architecture. Concurrently, Pini et al.^6^ highlighted important biological constraints of LNM, particularly regarding complex, non-linear connectivity changes following focal injury.

While robust statistical null models and biological realism are essential, we draw attention to a foundational, deterministic property that operates even under strictly linear connectivity assumptions at the single-patient level. Specifically, the spatial extent of a lesion does not merely introduce random noise; rather, it drives a systematic “bystander effect” rooted in how a lesion seed is operationalized. Whereas functional connectomes obey intrinsic functional parcellations, clinical lesions follow biological and pathological patterns—such as arterial territories, venous drainage, perilesional oedema, or infiltrative growth—that rarely align with functional boundaries.^7,8^ Consider a typical middle cerebral artery stroke causing expressive aphasia: while the symptom may be driven by damage to a focal critical node within Broca’s area, the vascular insult almost invariably encompasses adjacent insula, opercular cortex, and underlying white matter. Thus, a clinical lesion typically consists of a symptom-driving critical node surrounded by a variable volume of functionally unrelated, asymptomatic bystander tissue. Because standard LNM defines the seed by averaging connectivity across the entire binary lesion mask, the critical node’s specific connectivity signature is mechanically diluted by the mass of surrounding bystander tissue.

Using empirical clinical cohorts, individual-level machine learning with size-matched controls, simulated ground-truth networks, and synthetic spatial merges, we examine how lesion spatial extent shapes LNM representations. We demonstrate that as lesion volume increases, the inclusion of bystander tissue systematically overwrites focal connectivity signals, driving a mathematical convergence toward the connectome degree vector that poses fundamental questions for post-hoc statistical correction.

## Materials and Methods

### Data Sources and LNM Approximation

Lesion masks, cohort metadata, and the normative connectome data were obtained from the repository provided by van den Heuvel et al.^4^ (https://github.com/dutchconnectomelab/lesionnetworkmapping/). All network processing utilized the combined Schaefer-1000 (cortical) and Melbourne-54 (subcortical) parcellations, yielding 1054 total parcels in MNI152 space.

For each lesion, damaged voxels were mapped to the atlas to identify the set of injured parcels (P). The 1054-dimensional lesion network mapping (LNM) profile was approximated by averaging the normative resting-state functional connectivity rows corresponding to the damaged parcels, a linear approximation technique demonstrated by van den Heuvel et al.:

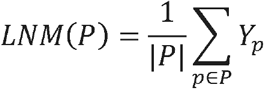

where Y is the connectivity matrix provided.^9^

### 9D Feature Space and Convergence Metric

Following the dimensionality reduction approach proposed in the literature^4^ to summarize these connectome profiles, we projected each 1054D LNM profile into a 9-dimensional functional subspace. This basis consists of: global connectivity degree, subcortical degree, functional connectivity to 4 independent resting-state network (RSN) modules, and the top 3 global connectivity gradients.

To quantify network map convergence, our target metric was the mean squared error (MSE). We measured the MSE between a lesion’s standardized 9D LNM profile and the connectome’s generic degree distribution vector (which constitutes the origin of the standardized feature space).

### Lesion Filtering and Inclusion Criteria

To ensure spatial stability, micro-lesions were excluded based on two criteria:

1. **Voxel threshold**: Lesions comprising , ≤ 100 total voxels in MNI space were removed.
2. **Parcel threshold**: A minimum affected-parcel count was strictly enforced so that lesions had a meaningful spatial network extent before generating aggregate connectome networks:

- for predominantly cortical and widely distributed lesion cohorts (Aphasia Recovery, LesyMap, OCD, and Schizophrenia, where mean cortical parcel overlap exceeded 84%; **Fig. 1b**), lesions intersecting , ≤ 5 parcels were discarded.
- For the Epilepsy cohort, anatomical mapping revealed a bimodal distribution containing a substantial subpopulation of purely subcortical lesions (0% cortical parcel overlap, reducing the cohort mean to 64.0% cortical; **Fig. 1b**). Because these subcortical lesions span only one to five contiguous parcels, a > 5 parcel threshold would artificially exclude this clinical subpopulation. A tailored minimum of ≥ 2 parcels was therefore employed for the Epilepsy dataset to exclude isolated single-parcel noise while preserving the representative clinical phenotype.

**Figure 1.**
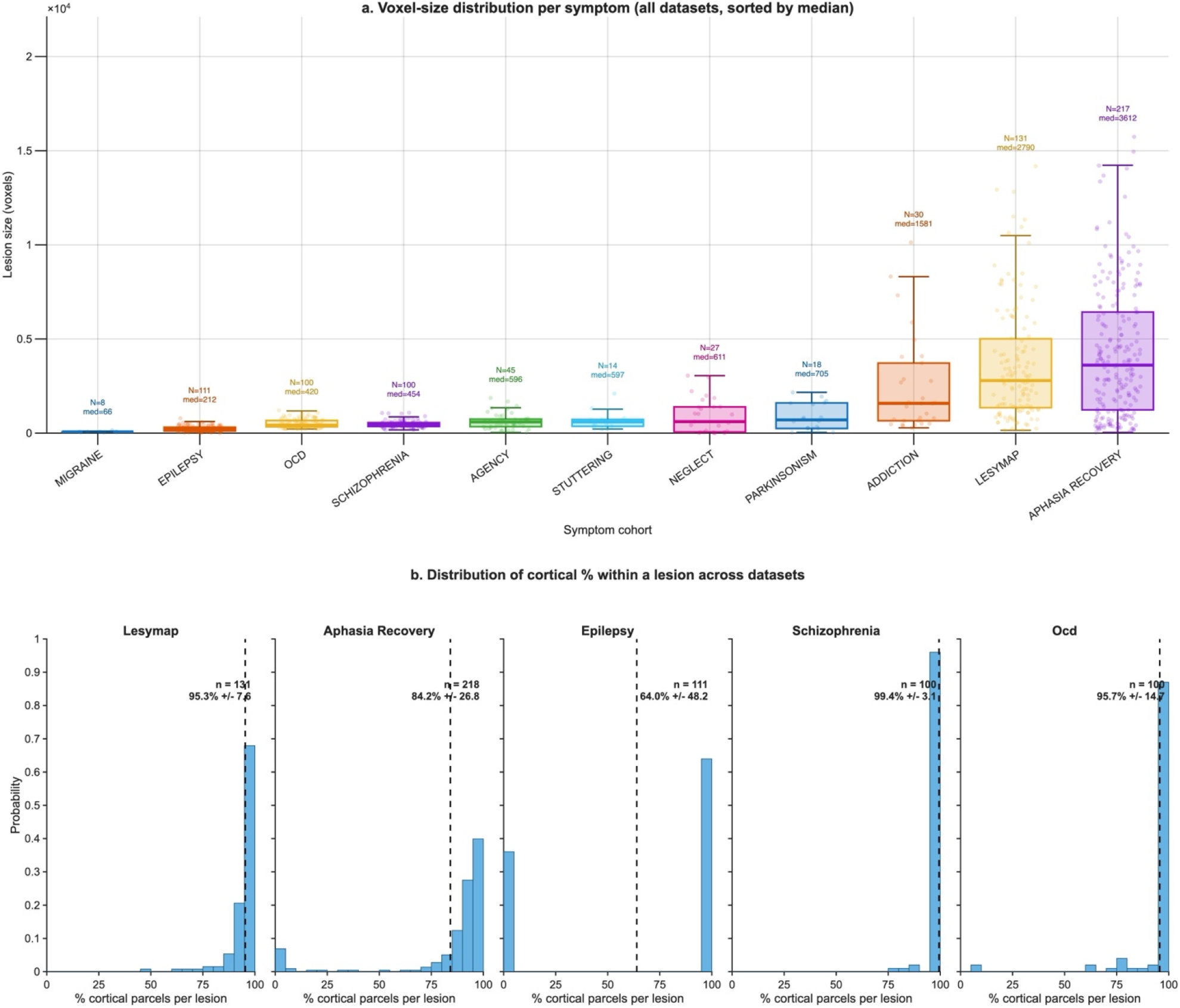
A. Variance of lesion size across cohorts: Lesions were taken from all cohorts available at https://github.com/dutchconnectomelab/lesionnetworkmapping/. Lesion were provided as binary masks. Lesion size was determined by number of voxels within the MNI152 space. Cohorts are sorted along the x-axis in ascending order of their median lesion size. For each distribution, the thick central line represents the median, while the lower and upper boundaries of the box indicate the 25th and 75th percentiles (the interquartile range, IQR), respectively. Whiskers extend to the most extreme data points that fall within 1.5 × IQR of the box edges. Individual patient lesions are overlaid. The total sample size (N) and exact median volume (med) are explicitly annotated above each respective cohort. **B. Distribution of cortical overlap across clinical lesion datasets.** Histograms display the percentage of cortical parcels intersected by each individual lesion across the five datasets. Vertical dashed lines denote the mean cortical percentage for each cohort. While lesions in the LesyMap, Aphasia Recovery, Schizophrenia, and OCD datasets are mostly cortical, the Epilepsy cohort contains a distinct, massive subpopulation of purely subcortical lesions (visible as the prominent spike at 0%).

Across the core comparative groups examined in this manuscript, parcel-thresholding resulted in 575 lesions reliably retained and 84 focal micro-lesions explicitly excluded.

### Power-law scaling analysis of lesion size and network collapse

The relationship between native lesion voxel size and mathematical distance to the degree (MSE) was modeled using ordinary least squares (OLS) regression in log-log space:

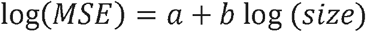

The resulting relationship is reported fundamentally as *MSE* ∝ *size*^*b*^ , alongside the explicit coefficient of determination (*R*^2^) to describe the convergence rate as lesion size scales.

### Classification of lesion size using Quadratic Discriminant Analysis

Utilizing the Aphasia Recovery and LesyMap datasets (the two largest cohorts demonstrating the most extreme volumetric variance, see Fig. 1A; Addiction dataset lacked sufficient lesions and other symptoms lacked size variance), lesions passing the voxel (>100) and parcel (> 5) filters were stratified exclusively by physical size (excluded: 1/130 LesyMap, 13/202 aphasia recovery). A 5-fold cross-validated Quadratic Discriminant Analysis (QDA) model was strictly trained to classify the top 25% largest lesions from the bottom 25% smallest lesions relying solely upon their 9D LNM network profiles. To verify statistical robustness (Fig. 2C), the empirically observed balanced accuracy was tested against a null distribution generated via 10,000 random, size-unaware label permutations.

**Figure 2.**
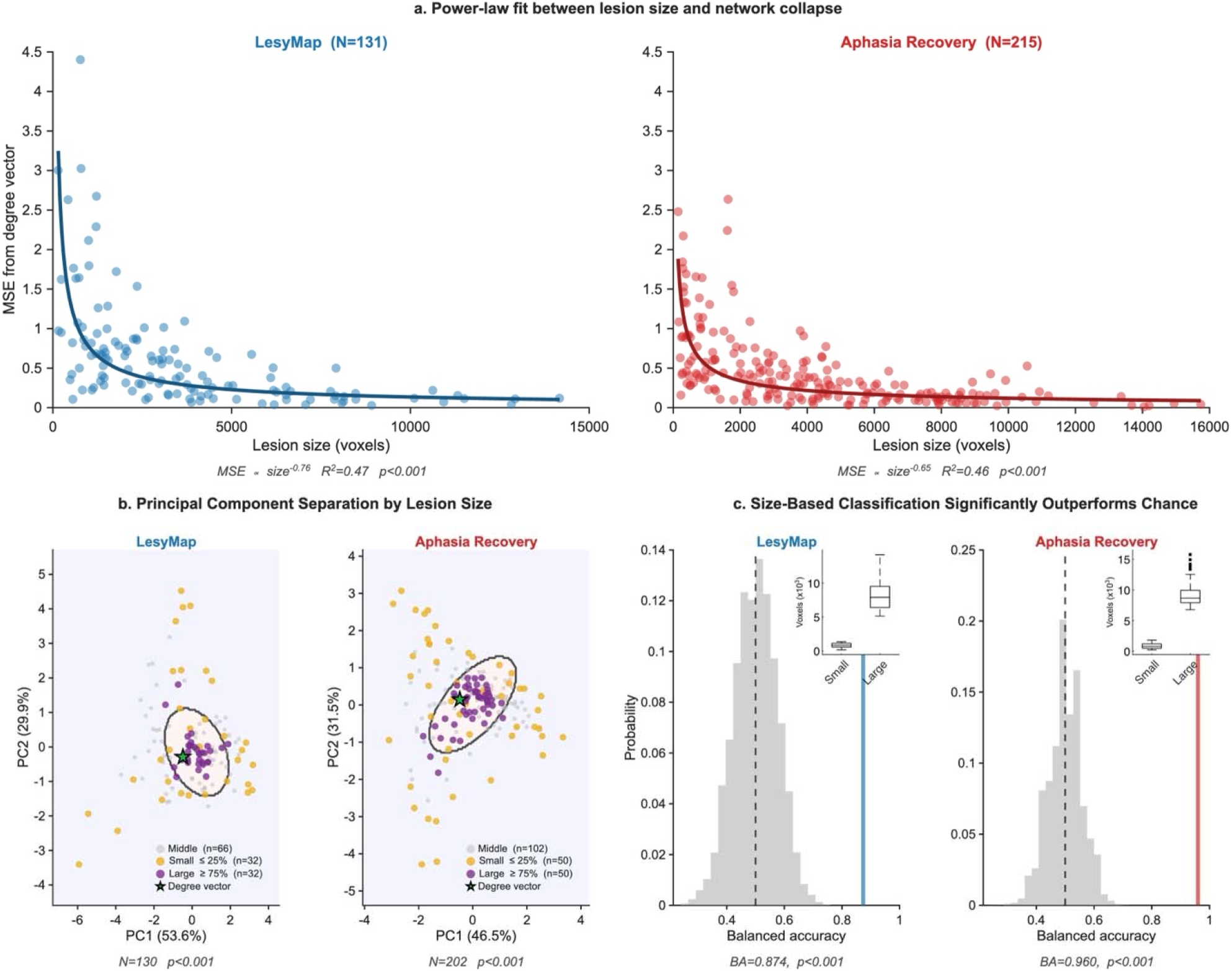
Size-driven convergence to the connectome degree. **A. Power-law fit between lesion size and network collapse.** The mean square error (MSE) of the 1054-dimensional lesion network map, relative to the connectome’s degree vector, decays as a strict power-law function of lesion size. This 1054d representation was calculated using van den Heuvel et al.’s4 linear approximation of LNM: parcellating lesions to the Melbourne-Schaefer parcellation and multiplying each binary vector by the precomputed connectome matrix. Size was measured by counting binary voxels in the original MNI152 space. **B. Principal component separation by lesion size**: Quadratic Discriminant Analysis (QDA) separating the top 25% largest from the bottom 25% smallest lesions (restricted to >100 voxels and >5 parcels for stability). Lesions were reduced from 1054D parcellation-space to a 9D LNM representation using van den Heuvel et al.’s dimensionality reduction, preserving almost all the explained variance (94.2% for LesyMap, 93.2% for Aphasia Recovery). The QDA boundary and data points are projected onto the two leading principal components for visualization. Large lesions form a tight Gaussian cluster around the degree vector, while small lesions spread across the rest of the subspace. **C. Size-based classification significantly outperforms chance:** Permutation testing of the large-versus-small discrimination (1B); across 10,000 permutations the large-versus-small partition yields the highest classification accuracy, indicating that LNM outputs are overwhelmingly driven by the size of the lesion. Insets display the underlying size distributions for the small and large lesion groups.

### Size-matched null modeling for individual-level classification

To prospectively assess whether individualized, disorder-specific machine learning classifiers inadvertently leverage systematic volume biases to achieve accuracy, an individual-level K-Nearest Neighbors (KNN) classifier was trained to separate target clinical cohorts from negative samples (pooled from overlapping datasets). The accuracy was calculated symmetrically under two independent conditions:

1. **Random null**: Negative samples were drawn natively at random from the aggregate remaining pool.
2. **Size-matched null**: Each specific positive clinical lesion was strictly paired to an immediate nearest-sized negative lesion from the remaining pool (computed greedily without replacement).

Differences in classifier discriminability formally clarify how much “specific” network topography signal is implicitly attributable to physical size variance.

### Simulation of spherical bystander lesions

To mechanically formalize the bystander effect, geometric spheres were simulated starting from the MNI centroid of all 1054 atlas parcels. At each discrete bounding radial step, all parcels whose true physical centroids fell within the expanding Euclidean radius were aggregated to operate as the “simulated lesion”.

### Construction of synthetic chimeric lesions

For *K* ranging from 2 to 10, *K* distinct lesions were randomly drawn from identical real-world patient cohorts such that they shared strictly zero overlapping parcels. These natively isolated parcel sets were systematically united to form a *K*-way “chimeric” lesion.

## Results

### Empirical lesion volume drives power-law convergence to the connectome degree

The collapse onto the degree vector is not merely an artifact of group averaging across patients, nor is it a statistical false positive that can be permuted away. It arises deterministically within a single patient as the lesion grows. As more bystander regions are incorporated into the unitary seed, the lesion’s specific connectivity profile is mechanically washed out, forcing the output map toward generic connectome hubs. We quantified this effect in the two datasets with the largest variance in lesion volume amongst datasets included in van den Heuvel et al.’s original data that also has n > 50 subjects (Aphasia recovery^10^ and LesyMap^11^; lesions ranging from ∼100 to ∼15,000 voxels). The mean squared error (MSE) between a lesion’s LNM representation and the degree vector decreases with lesion size following a power law in both datasets (LesyMap, *MSE* ∝ *size* ^− 0.76^,*R*^2^ = 0.47, p < 0.001; Aphasia Recovery, *MSE* ∝ *size*^− 0.65^ , *R*^2^ = 0.46, p < 0.001, **Fig. 2a**). We also trained a quadratic discriminant analysis (QDA) classifier to distinguish the largest 25% and smallest 25% lesions within each of the datasets, using van den Heuvel et al.’s 9D dense representation of lesions, and used permutation-based (10000 permutations) testing to assess significance. In both datasets, the classifier had the best accuracy amongst permutations (LesyMap, accuracy = 87.4%; Aphasia Recovery, accuracy = 96.0%; **Fig 2b,c**), demonstrating that a lesion’s spatial extent fundamentally dictates its network profile.

### Controlling for lesion size accounts for individual-level disorder classification

We next examined whether this volumetric scaling effect accounts for the apparent success of individual-level machine learning approaches in preserving clinical specificity. A recent proposal^5^ has suggested that shifting from group-level maps to individual-level analyses can isolate disorder-specific signals from generic connectome architecture, citing our prior work^12^ applying a K-Nearest Neighbors (KNN) classifier to reduced-dimensionality LNM representations. To test whether these individual-level representations are similarly confounded by bystander volume, we re-evaluated our classifier using a strict size-matched null model, pairing each clinical lesion to the closest-sized lesion drawn from other clinical cohorts. Controlling for lesion size drove classification accuracy for previously separable cohorts back toward chance levels (Permutation p-values, random → size-matched null: Aphasia Recovery < 0.001 → 0.06; OCD 0.005 → 0.42; Epilepsy < 0.001 → 0.03; Schizophrenia 0.16 → 0.28, Fig. 2a). Thus, the across-lesion variability that initially appeared to capture distinct clinical signatures primarily reflects systematic differences in lesion size distributions across cohorts (**Fig. 1a**). The modest residual signal observed in epilepsy (56.8 %, *p* = 0.03) likely reflects a broad anatomical dichotomy between cortical and subcortical structures, as a substantial subpopulation of lesions in this cohort is uniquely restricted to subcortical nuclei (**Fig. 1b**).

### Mathematical basis of bystander dilution in simulated and chimeric lesions

To verify that this convergence is a direct mathematical consequence of spatial extent rather than cohort-specific confounds, we simulated spherical bystander lesions originating from the centroid of every parcel in the Schaefer-Melbourne 1054-region atlas. As the simulated lesions expanded symmetrically from their original ground-truth seeds to incorporate adjacent tissue, the resulting LNM profile progressively diverged from the true seed connectivity and converged toward the connectome degree vector, regardless of the anatomical origin (**Fig. 3b**).

**Figure 3.**
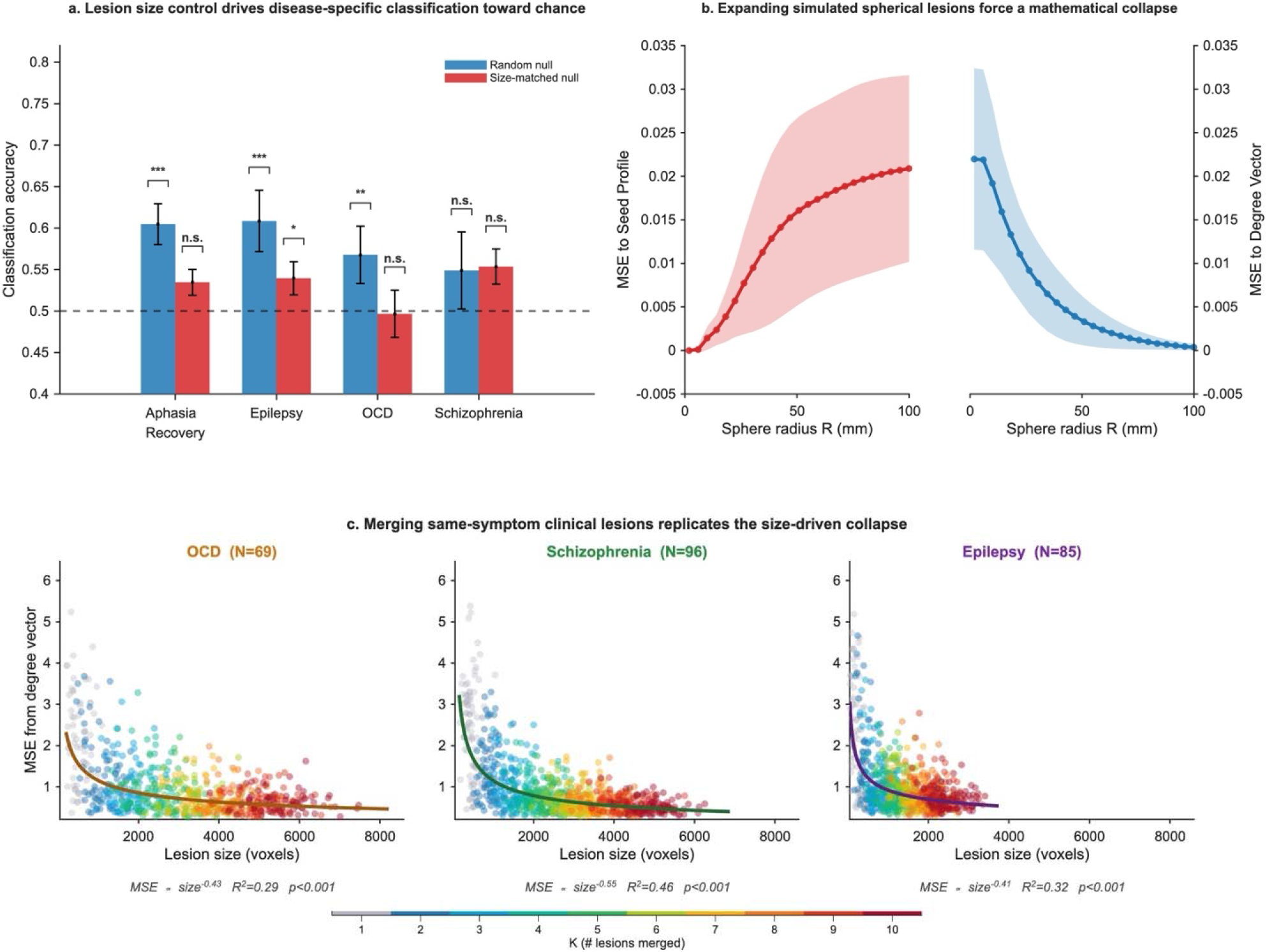
Size-driven collapse in real, simulated, and synthetic networks. A. Lesion size control drives disease-specific classification toward chance: Accuracy separating each clinical lesion set from a null set of other-disorder lesions, using an individual-level classifier on 9D LNM features (5-fold cross-validation; mean ± SD). Blue: null sampled at random, results taken from Edelman, Elias & Arzy;^12^ red: null size-matched to the clinical set (each patient paired to the nearest-sizedother-disorder lesion, without replacement). Permutation p-values (label shuffling), random → size-matched null: Aphasia < 0.001 → n.s. (0.06); OCD 0.005 → n.s. (0.42); Schizophrenia n.s. (0.16) → n.s. (0.28); Epilepsy < 0.001 → 0.03. Size-matching substantially reduces the disorder signal, leaving only a weak residual in epilepsy. B. Expanding simulated spherical lesions force a mathematical collapse: To mechanically isolate the effect of bystander volume from spatial location, we simulated spherical lesions originating from the MNI centroid of every individual parcel (N = 1054) as independent ground-truth seeds. Starting from the isolated seed parcel (*R* = 2 mm), the lesion radius (*R*) was iteratively expanded to incorporate adjacent bystander parcels. The LNM output at each step was calculated as the mean connectivity profile of all encapsulated parcels. As the lesion expands spherically, the physical inclusion of bystander tissue deterministically forces the LNM output to diverge from the original seed’s profile (red curve, left) and systematically collapse toward the generic degree vector of the connectome (blue curve, right). Solid lines denote the mean squared error (MSE) across all 1054 simulations; shaded regions represent ± 1 SD. C. Merging same-symptom clinical lesions replicates the size-driven collapse: Power-law fit between synthetic lesion size and MSE to the degree vector, for lesions constructed by merging *K* non-overlapping same-symptom clinical lesions (*K* = 2 − 10). The bottom colorbar indicates the number of merged lesions (*K*). The same power-law link between lesion size and the degree vector re-emerges across all three distinct clinical cohorts.

Mathematically, this power-law scaling follows directly from the linear odemonstrating that a lesion′s spatial sperationalization of LNM. Because a lesion’s network profile is computed as the sample mean of the connectivity rows corresponding to its constituent spatial units, and the global mean across all rows of the connectome matrix defines the degree vector, an expanding lesion acts as an growing sample drawn from the connectome. Under the Central Limit Theorem, the mean squared error (MSE) between a sample mean of size *N* and the population mean decays as a power law (MSE ∝ *N* ^− 1^) for independent draws. In the brain, because spatially contiguous parcels exhibit strong local autocorrelation rather than statistical independence, the effective degrees of freedom grow more slowly than raw voxel or parcel counts, yielding the empirically observed sub-unit power-law exponents (MSE ∝ size ^− *b*^, with *b* ≈ 0.41--0.76).

A natural objection is that these simulated spheres do not reflect true biological damage, and that in real clinical data, lesion size is inextricably confounded with specific lesion locations. To isolate size while maintaining empirical lesion topography, we constructed synthetic lesions in the three remaining large datasets (N > 50) that have low native size variance comprising patients with epilepsy,^13^ schizophrenia^14^ and obsessive-compulsive disorder (OCD).^15^ For each K from 2 to 10, we merged K non-overlapping lesions drawn from the same dataset. Such unions are biologically reasonable: if a lesion at A and a lesion at B independently produce a symptom X, their union is unlikely to fall outside the symptom distribution. The exact same power-law relationship between (synthetic) lesion size and distance to the degree vector re-emerged (OCD, *MSE* ∝ *size*^− 0.43^,*R*^2^ = 0.29; Schizophrenia, *MSE* ∝ *size* ^− 0.55^,*R*^2^ = 0.46; Epilepsy, *MSE* ∝ *size* ^− 0.4l^, *R*^2^ = 0.32; all three *p* < 0.001, **Fig. 3c**), confirming that spatial extent and bystander dilution - not merely a correlation with location - mechanically overwrite specific clinical signals, even when strictly constrained by empirical lesion topography.

## Discussion

This real, synthetic and simulated collapse highlights a fundamental limitation in relying solely on post-hoc statistical corrections. It may be assumed that a spatial null model could correctly render the maps of these large synthetic lesions non-significant, thereby protecting against false positives. However, this conflates a failure of inference with a failure of measurement. Because dilution occurs at the seed level - physically averaging specific connectivity profiles out of existence before a map is generated - the resulting output might be dominated by the volume of bystander tissue. Consequently, even when an individual map survives a spatial null model and is deemed statistically significant, its biological interpretation remains ambiguous. It becomes nearly impossible to untangle whether the yielded network genuinely reflects the specific causal circuitry of the symptom, or simply a mixture heavily contaminated by the network profile of asymptomatic bystander areas.

It is well established that traditional lesion mapping techniques (such as voxel-based lesion-symptom mapping^16^) are highly vulnerable to anatomical and vascular biases, and to factors like size or shape of the lesion.^7,8^ However, while in classical VLSM these biological constraints primarily confound post-hoc statistical inference due to a violation of statistical assumption, in traditional LNM they create a foundational artifact that invalidates the measurement itself, by forcing the entire lesion mask to define the connectivity profile. Larger or multi-focal lesions mathematically converge on the connectome degree, therefore lesion size can drive LNM results entirely on its own. This artifact might be even stronger in newer variations of LNM that extend the notion of a “lesion” to multi-focal, distributed spheres like white-matter tracts^17^ or continuous thresholded neurodegeneration markers such as atrophy^18^ or tau.^19^ One proposed workaround is to restrict the seed to the lesion’s centroid;^20^ however, because biological damage does not follow functional topography, there is no guarantee that the anatomical center of a lesion aligns with the critical functional node driving the symptom.

Importantly, while our analyses build upon a parcellated linear approximation of LNM, the bystander effect applies equally to classical voxel-wise LNM pipelines. In standard voxel-level LNM, the resting-state BOLD time series is first averaged across all voxels within the binary lesion mask in each normative subject prior to computing whole-brain correlations and group-level *t*-statistics. Because Pearson correlation and expectation are linear operations up to a scaling factor, averaging thousands of voxel time series across a large lesion mask mechanically attenuates focal variance at the time-series level. As the volume of co-averaged bystander voxels grows, the mean seed time series becomes increasingly dominated by shared global and high-degree fluctuations, driving the resulting voxel-wise connectivity map toward the same generic degree architecture before any statistical thresholding is applied.

Understanding the link between focal damage and distributed network dysfunction remains one of the most compelling goals in clinical cognitive neuroscience, and the conceptual framework of LNM has rightly shifted the field toward a circuit-level perspective. Realizing the full diagnostic and therapeutic potential of this paradigm does not require abandoning connectome-based mapping but rather refining how lesion seeds are defined. Future methodological advances should focus on distilling the functionally causal lesion core from the raw anatomical mass of surrounding bystander tissue—for example, by decomposing heterogeneous lesion masks into functionally homogeneous sub-parcels prior to projection. Addressing this spatial dilution at the seed level will provide a rigorous foundation for the next generation of lesion-based network models.

## Data Availability

The data that support the findings of this study are publicly available. Lesion masks, cohort metadata, and code are available from the repository provided by van den Heuvel et al. (https://github.com/dutchconnectomelab/lesionnetworkmapping/).

## Data Availability

The data that support the findings of this study are publicly available. Lesion masks, cohort metadata, and code are available from the repository provided by van den Heuvel et al. (https://github.com/dutchconnectomelab/lesionnetworkmapping/). The normative connectome data are available via the Harvard Dataverse (https://doi.org/10.7910/DVN/ILXIKS).

